# Prevalence of Zoonotic Parasites in Non-Ruminants and Humans in the Wa West District of Ghana: A One Health Perspective

**DOI:** 10.1101/2025.05.27.25328388

**Authors:** Joshua Kpieonuma Zineyele, Denis DekugmenYar, Nana Yaa Awua-Boateng, Ebenezer Assoah

## Abstract

This study assessed human and domesticated non-ruminant interaction and the risks of parasitic zoonosis in the One-Health Concept in the Wa West District of Ghana. Using a descriptive cross-sectional design, 286 faecal samples were collected, 221 from non-ruminants and 65 humans. Using a structured questionnaire, participants were interviewed face-to-face, while faecal samples of animals and humans were collected and examined using concentration techniques. The majority and 26 and 34 years old. Most (69.0%) participants raised animals using an extensive system, and 50.0% did not routinely inspect or monitor their animals’ health. The prevalence of intestinal parasites among the non-ruminants and humans was 60.60% and 67.69%, respectively. Of all the faecal samples examined, 21 parasite species were recovered, of which 14 were isolated from humans. *Ancylostoma caninum*, *Cryptosporidium* sp., and *Strongyloides* sp., were the predominant parasites in the non-ruminants. In humans, *Bunostomum* sp.*, Cryptosporidium* sp.*, Emeria* sp.*, Strongylus* sp.*, and Toxocara cati* were the most prevalent parasites, each with a rate of 7.70%. *Ancylostoma caninum, Toxocara canis, Toxocaris leonina, and Trichuris vulpis* were recovered from dogs, their specific host. However, *Cryptosporidium* sp.*, Ascaris* sp.*, Paragonimus kellicotti,* and *Strongyloides* sp., whose specific host is man, were also recovered in non-ruminants. This study recorded a high prevalence of multiple parasitic infections with cross-transmission of species between non-ruminants and humans with zoonotic potential in the Wa West District. The cross-transmission of parasite species between host-specific non-ruminants and humans is linked to their close interaction and sharing of a limited space. The outcome of this study has dire implications for animal and human health and calls for implementing the One-Health Concept to curb the outbreak of parasitic zoonosis in this region.

**Author Summary:** Zoonotic parasitic infections pose a significant public health challenge, especially in rural communities where close contact with domestic animals is common. This study, conducted in the Wa West District of Ghana, applied a One Health approach to investigate the prevalence and types of intestinal parasites in humans and domesticated non-ruminant animals such as dogs, pigs, poultry, and cats. A total of 286 fecal samples (221 from animals and 65 from humans) were analyzed using concentration techniques, and 21 parasite species were identified, with notable overlap between those found in humans and animals. The study revealed high infection rates: 60.60% in animals and 67.69% in humans. Several parasites, including *Cryptosporidium* spp. and *Strongyloides* spp., were found in both hosts, indicating active cross-transmission. These findings emphasize the public health risks associated with poor animal husbandry practices, lack of routine veterinary care, and shared based One Health interventions that promote animal and human health simultaneously through coordinated surveillance, improved hygiene, and education in rural Ghana and similar settings.

## 1. Introduction

Globally, zoonotic diseases are increasing and constitute a significant public health threat in recent times [1]. Over 60.0% of human diseases are infectious, with approximately 75.0% originating from animal sources [2]. However, the dynamics of human-domestic animal interaction play a pivotal role in the spillover of infectious pathogens of zoonotic importance [3]. More than 2.5 billion cases of zoonosis occur each year, with about 2.7 million deaths worldwide. [4]. Moreover, emerging and re-emerging zoonoses such as COVID-19, Monkey pox disease, Ebola virus disease, and Avian influenza highlight the ongoing threat posed by pathogens originating from animal reservoirs [5].

In Africa, the interaction between humans and domestic animals is commonplace and is influenced by several factors, including cultural, environmental, and socio-economic [6]. The African continent has witnessed about a 63.0% increase in cases of zoonotic diseases in the last decades[7]. Over 70.0% of the African population, however, relies on backyard animals rearing for their livelihoods and often coexist closely with these domestic animals [8]. However, the proximity and interaction between humans, domestic animals, and wildlife increases the potential for the transmission of pathogens of zoonotic importance [9].

Moreover, most of the annual millions of deaths in Sub-Saharan Africa are due to limited access to healthcare services, close contact with animals, and environmental conditions conducive to the transmission of infectious diseases [10]. Nearly 25.0% and 70.0% of these diseases are zoonotic which are linked to domestic and wild animals, respectively [11]. This, therefore, highlights the critical role domestic and wild animals contribute to the transmission of zoonotic pathogens of public health importance [12].

About 70% of Ghana’s population engages in agriculture, including backyard animal farms with high human-animal interactions, which makes them vulnerable to zoonotic diseases [13]. Reports in Ghana show rabies is endemic due to the failure to vaccinate dogs [14], resulting in human mortality estimated at 1000 annually [15], especially in rural areas [16]. Furthermore, 6.7% and 92.0% of brucellosis and toxoplasmosis were recorded in cattle and humans, respectively, in rural Ghana [17]. In addition, humans, domestic animals, and wildlife animals’ intimate interaction is more prevalent in rural areas, creating a unique interface for zoonosis transmission [18]. This highlights the One-Health Concept, which indicates the interconnection and interplay between human and animal health and well-being and their interaction with the environment.

In Ghana, several studies focused on different zoonotic parasites, including the knowledge of zoonoses [19], tick-born parasites in livestock [20], giardiasis, and helminths in cattle and dogs [21]. However, these studies did not examine the interaction between humans and domestic animals in rural communities and the potential of parasitic zoonoses. Meanwhile, there is a high potential risk of infectious pathogens spilling from animals to humans through the environment. Besides, limited literature on parasitic zoonosis exists in the Wa West District. Moreover, in this district, most households are engaged in backyard farms and practicing free-range animal husbandry. There are reports of zoonosis in this study area with uncertain sources [22]. This has become imperative and thus calls for an urgent need to assess the interaction and coexistence between humans and non-ruminants within household settings and the risk of parasitic zoonosis. This is crucial for informing evidence-based interventions and policy considerations [23]. Therefore, this study sought to assess the prevalence of parasites in non-ruminants and humans, their coexistence in household settings, and the risk of parasitic zoonosis in the One-Health Concept in the Wa West District of Ghana.

## 2. Materials and Methods

### Study Area

This study was conducted in the Upper West region of Ghana, with Wa as its administrative capital (**Figure 1**). The Upper West region is one of the 16 regions in Ghana, and it is located in the north-western part of the country, with latitude 9.8^0^-11.0^0^ North and longitude 1.6^0^-3.0 West. Specifically, the study was conducted in the Wa West district, located in the western part of the region. It shares boundaries to the West with the Ivory Coast, North with the Nadowli/Kaleo district, East with Wa Municipality, and South with the Sawla/Tuna/Kalba district. Wechiau is the administrative capital, and the district has a total population of 96,957, comprising 45,880 males and 51,077 females [24]. They are predominantly farmers with an average household ratio of 5.4, the highest in the region. Most households have backyard livestock, including poultry, pigs, sheep, goats, and cattle [25]. Cats and dogs are also raised for hunting, security, and other purposes. The district lies between Latitude 9.82556 degrees North and Longitude –2.66927 degrees West. The study sites in the district are indicated on a map in Figure 1 with circles: Berinyasi, Bultuo, Dabo, Ga, Grungu, Kendeo, Ponyenitanga, Siiroo, Tanvaare, Varinpere, and Vieri.

**Figure 1:**
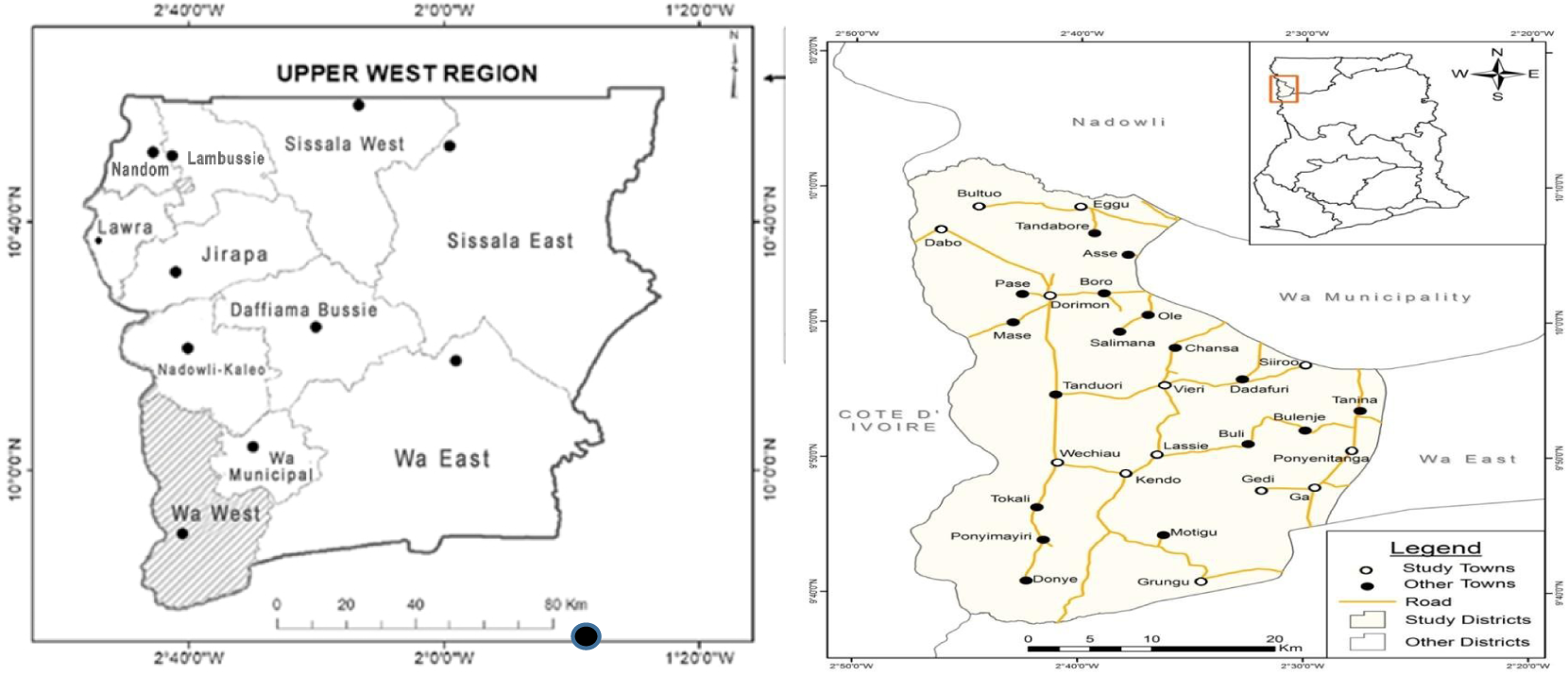
Map of the Upper West Region. Source: Ghana Statistical Service (GSS)

### Research design

In a One-Health Concept, a descriptive cross-sectional design assessed human-domesticated animal interactions and the risk of parasitic zoonosis. This design was preferred as it measured different variables and analyzed the relationship between exposures and outcomes.

### The population of the study

The study population included all persons between the ages of 6 and 50 years and non-ruminants within the Wa West District, such as dogs, pigs, cats, rabbits, and poultry.

### Sampling techniques

This study employed a multistage sampling technique. First, the study area was clustered into five clusters. Second, two communities were randomly selected per cluster. Third, a purposive technique was used to select households with backyard farm animals. One household member was randomly chosen, and a convenient method was used for collecting faecal samples.

### Research instrument

A structured questionnaire was developed into five sections: demographics and house members and characteristics, number and types of domesticated animals and levels of confinement, animal husbandry practices, veterinary services, and knowledge of zoonosis. Also, a checklist was developed and coupled with observation to capture the number and types of backyard farm animals, levels of confinement, animal-human interaction, husbandry practices, and litter of animal droppings in the household compound. Lastly, a laboratory tool was developed for faecal sample collection and microscopy examination results.

### Data collection procedure

The English questionnaire was translated into local dialects, including Dagaare, Waale, and Brefor. Face-to-face interviews were employed to elicit answers from the study participants using questionnaires. A checklist was used to collect data on the types of animals, numbers, and levels of confinement from each household.

### Faecal Sample Collection

Faecal samples from domesticated animals were collected directly from the animal’s rectum or freshly voided. About 80g of fresh animal and human faecal samples were collected into clean, transparent, well-labelled containers. Animal and human data such as age and sex were also collected.

### Laboratory Methods

The faecal samples were analyzed using concentration techniques (sedimentation and floatation), wet mount, and Acid-Fast Stain. The centrifugal flotation method using different solutions (Sodium chloride, Sodium nitrate, zinc sulfate, and formal ether) was used to recover the parasite ova, oocysts, or larvae based on their specific gravity in faecal samples of domestic animals, as previously reported [26]. The samples were then microscopically examined with 10X and 40X objectives. As reported earlier, the McMaster technique was also employed to count the total number of parasites per gram of each faecal sample [27]. Zinc sulfate solution was used to recover *Ancylostoma* sp. and *Toxocara cati*. Acid-fast staining was used to aid with the microscopic examination of *Cryptosporidium* sp. The different parasites and species were identified using morphological characteristics and staining techniques as previously described [14].

### Data Management and Analysis

The statistical tool of R software was used to calculate the one-way ANOVA for the raw data. A p-value of 0.05 significant level was calculated using a confidence interval (C.I) of 95% to compare the prevalence of parasites between humans and domesticated animals. Descriptive statistics were employed to determine the frequency and percentages of the cross-transmission of host-specific parasites. Bar graphs and tables were used to assess the prevalence of parasites in humans and domestic animals and their association with socio-demographic and environmental factors.

### Ethical Review and Clearance

Ethical clearance was obtained from the Committee on Human Research, Publications, and Ethics at the Kwame Nkrumah University of Science and Technology (Ref: CHRPE/AP/851/23). Prior to sample collection, written permission was obtained from the Wa West District Assembly. Each participant received a detailed verbal explanation of the study’s objectives, procedures, potential risks, and benefits. They were also informed of their right to participate voluntarily and their freedom to withdraw from the study at any time without any consequences. After the briefing, participants who agreed to take part signed a written informed consent form, confirming their voluntary participation.

## 3. Results

### Socio-Demographic Characteristics

**Table 1** shows that 64.0% and 36.0% of the household heads were males and females, respectively. Also, 42% of the respondents were between 41 and 49, and 6.0% were between 26 and 34 years. Most (83.0%) respondents were married; 7.0%, 9.0%, and 1.0% were single, divorced, and windowed, respectively. Most participants (81.0%) had no formal education, whereas 19.0% had at least a primary education. The majority (64.0%) of the respondents were Christians, 22.0% were Muslims, and 14.0% were Traditionalists. Also, 92.0% of the respondents were farmers, 6.0% were teachers, and 2.0% were petty traders. Most (69.0%) households had a large family of more than ten members, while 42.0% had between 3 and 4 children (See **Table 1**).

**Table 1:** Socio-Demographic Characteristics of the Respondents.

| Variable | Frequency | Percentage (%) |
| --- | --- | --- |
| **Gender** |  |  |
| Male | 64 | 64.0 |
| Female | 36 | 36.0 |
| **Age Range** |  |  |
| 18-25 | 15 | 15.0 |
| 26-34 | 6 | 6.0 |
| 35-40 | 17 | 17.0 |
| 41-49 | 42 | 42.0 |
| 50-above | 19 | 19.0 |
| **Educational Status** |  |  |
| Literate | 19 | 19.0 |
| Illiterate | 81 | 81.0 |
| **Employment status** |  |  |
| Employed | 6 | 6.0 |
| Unemployed | 91 | 91.0 |
| Self-employed | 3 | 3.0 |
| **Occupation** |  |  |
| Farmer | 92 | 92.0 |
| Teachers | 6 | 6.0 |
| Petty Trader | 2 | 2.0 |
| **Family Size** |  |  |
| 1-3 | 4 | 4.0 |
| 4-6 | 6 | 6.0 |
| 7-9 | 21 | 21.0 |
| ≥10 | 69 | 69.0 |
| **Children in Household** |  |  |
| 1-2 | 20 | 20.0 |
| 3-4 | 42 | 42.0 |
| 5-6 | 25 | 25.0 |
| ≥7 | 12 | 12.0 |

Source: Fieldwork

### Animals and Husbandry Practice

**Table 2** shows that all the respondents had animals in their homes, using either an extensive system (69.0%) or a semi-intensive system (31.0%) for rearing them. However, 49.0% of respondents do not routinely monitor animals, whereas 46.0% and 5.0% are regularly and sometimes monitored for diseases. Most respondents (98.0%) indicated that their animals were not confined, and 66.0% did not feed them at home, whereas 21.0% and 13.0% fed their animals daily and occasionally. Concerning the type of animals reared, 96.0% kept poultry, 91.0% kept goats, while 63.0%, 54.0%, and 51.0% kept sheep, dogs, and pigs, respectively. Also, 49.0% and 17.0% kept cats and cattle, respectively, as shown in **Table 2**. The majority (63.0%) of the respondents raised goats, while 14.0%, 11.0%, 10.0%, and 2.0% said sheep, cattle, poultry, and pigs for commercial purposes respectively.

**Table 2:** Animals and Husbandry Practice.

| Variables | Frequency | Percentage (%) | p-value |
| --- | --- | --- | --- |
| **Present of animals in the household** |  |  |  |
| Yes | 100 | 100.0 | <0.001 |
| **Number of animals you own** |  |  |  |
| 1-19 | 9 | 9.0 | <0.001 |
| 20-39 | 12 | 12.0 |  |
| 40-69 | 3 | 3.0 |  |
| 70->80 | 76 | 76.0 |  |
| **Routine disease monitoring of animals** |  |  |  |
| Yes | 46 | 46.0 | <0.001 |
| No | 49 | 49.0 |  |
| Sometimes | 5 | 5.0 |  |
| **Animals are allowed to roam about** |  |  |  |
| Yes | 98 | 98.0 | <0.001 |
| No | 2 | 2.0 |  |
| **Form of rearing/raising the animals.** |  |  |  |
| Semi-intensive | 31 | 31.0 | <0.001 |
| Extensive | 69 | 69.0 |  |
| **Rear animals for commercial purposes** |  |  |  |
| Goats | 63 | 63.0 | <0.001 |
| Sheep | 14 | 14.0 |  |
| Cattle | 10 | 10.0 |  |
| Poultry | 11 | 11.0 |  |
| Pigs | 2 | 2.0 |  |
| **Feed animals daily** |  |  |  |
| Yes | 21 | 21.0 | <0.001 |
| No | 66 | 66.0 |  |
| Sometime | 13 | 13.0 |  |
| **Animals you own** |  |  |  |
| Poultry | 96 | 96.0 | <0.001 |
| Goat | 91 | 91.0 |  |
| Dogs | 54 | 54.0 |  |
| Cats | 49 | 49.0 |  |
| Sheep | 63 | 63.0 |  |
| Cattle | 17 | 17.0 |  |
| Pigs | 51 | 51.0 |  |

*Source: Fieldwork*

**Table 3** shows an overall parasite prevalence of 60.60% in the study communities, with the Kendeo community recording a prevalence of 90.50%. Meanwhile, dogs, cats, and poultry had the highest parasite prevalence of 68.20%, 66.70%, and 28.30%, respectively. Ga community recorded 89.20% parasite prevalence, with low rates for poultry (0.21%) and dogs (0.054%). The Ponyenitanga had 88.1% parasite prevalence, with the highest rate in poultry (23.8%) and 7.14% and 4.76% rates in pigs and dogs. The parasitic infection rate among the animals in the Grungu community was 73.70%, of which poultry (31.60%) and cats (1.75%) recorded higher rates. The Yeleyiri community had a 70.30%. Berinyasi, 65.80%, 65.40% in Siiroo, 62.20% in the Tanvaare community, and the Vieri community, 7.0%.

**Table 3:** Parasitic Infection Rates among Communities and Animals.

| Animal Type | Cats<br>n (%) | Dogs<br>n (%) | Ducks<br>n (%) | Guinea<br>n (%) | Pigs<br>n (%) | Poultry<br>n (%) | Total<br>n (%) | P-value |
| --- | --- | --- | --- | --- | --- | --- | --- | --- |
| **Community** |  |  |  |  |  |  |  |  |
| **Berinyasi** | 1(2.63) | 5(13.15) | 0 | 0 | 3(7.89) | 4(10.5) | **13 (65.8)** | 0.0002 |
| **Ga** | 0 | 2(0.054) | 0 | 0 | 0 | 8(0.21) | **10 (89.2)** | <0.001 |
| **Grungu** | 1(1.75) | 2(3.51) | 0 | 0 | 5(8.77) | 18(31.6) | **26 (73.7)** | <0.001 |
| **Kendeo** | 0 | 2(3.77) | 0 | 0 | 0 | 15(28.3) | **17 (90.5)** | <0.001 |
| **Ponyenitanga** | 0 | 2(4.76) | 1(2.38) | 0 | 3(7.14) | 10(23.8) | **16 (88.1)** | <0.001 |
| **Siiroo** | 0 | 0 | 1(1.92) | 0 | 0 | 12(23.1) | **13 (65.4)** | <0.001 |
| **Tanvaare** | 0 | 1(2.7) | 0 | 0 | 2(5.41) | 8(21.62) | **11 (62.2)** | <0.001 |
| **Varinperi** | 0 | 0 | 0 | 0 | 1(3.57) | 2(7.14) | **03 (42.8)** | 0.004 |
| **Vieri** | 0 | 0 | 0 | 1(3.33) | 0 | 11(36.7) | **12 (70.0)** | 0.054 |
| **Yeleyiri** | 0 | 1(1.56) | 0 | 0 | 11(17.18) | 1(1.56) | **13 (70.3)** | 0.065 |
| **Total** | **2(66.7)** | **15(68.2)** | **2(40)** | **1(50)** | **25(65.8)** | **89(59.3)** | **134(60.6)** | **<0.001** |

### Prevalence of Parasites among Study Sites and Domesticated Animals

Figure 2 shows the parasite rates among the study sites and the seven non-ruminant domestic animals. The prevalence of parasites in dogs was 68.20%, 66.70% in cats, 65.80% in pigs, 59.30% in poultry, 50.0% in guinea fowls, and 40.0% in ducks, with females recording 77.30%.

**Figure 2:**
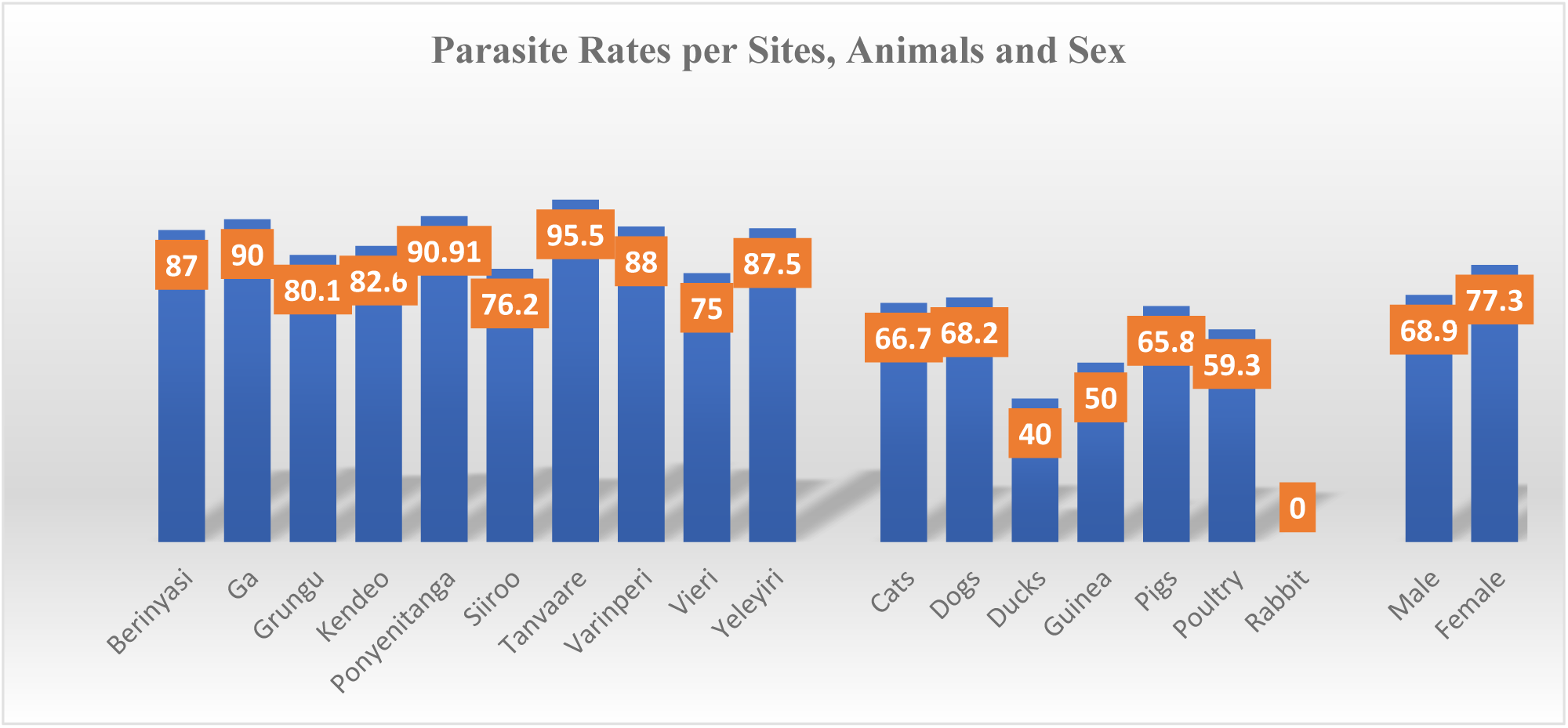
Prevalence of Parasites among Study Sites and Domesticated Animals.

### Prevalence of Parasite Isolates among Domesticated Animals

**Table 4** shows that in guinea fowls, *Ancylostoma caninum, Heteraki galliae, and Isospora* sp. were recovered from the samples. Also, *Ancylostoma caninum, Ascaridia* sp*., Bunostomum* sp*., Echinococcus* sp.*, Meullerius capilaris, Moniezia benedeni, Trichuris vulpis, and Isospora* sp. were the parasites isolated from pigs. *Heteraki galliae, Emeria* sp*., Paragonimus kellicotti, Strongyloides* sp*., Toxocara cati, Trichuris vulpis, and Isospora* sp. were identified in poultry. In the study area, rabbits were also noted to be infected with *Ancylostoma caninum, Nanophyetus* sp., and *Cryptosporidium* sp., with one infection each.

**Table 4:** Prevalence of parasites among non-ruminants and humans.

| **Parasite** | **Cats** | **Dogs** | **Ducks** | **Guinea fowls** | **Pigs** | **Poultry** | **Rabbit** | **Total** | **Percentage (%)** | **P-value** |
| --- | --- | --- | --- | --- | --- | --- | --- | --- | --- | --- |
| *Ancylostoma caninum* | 1 | 15 | 1 | 1 | 12 | 0 | 1 | 31 | 25.0 | <0.001 |
| *Ascaridia* sp. | 0 | 1 | 0 | 0 | 2 | 0 | 0 | 3 | 2.4 | 0.0086 |
| *Bunostomum* sp. | 0 | 1 | 0 | 0 | 6 | 0 | 0 | 7 | 6.0 | <0.001 |
| *Echinococcus* sp. | 0 | 0 | 0 | 0 | 1 | 0 | 0 | 1 | 0.8 | 0.245 |
| *Fasciola* sp. | 1 | 0 | 0 | 0 | 0 | 0 | 0 | 1 | 0.8 | <0.001 |
| *Heteraki galliae* | 0 | 0 | 0 | 1 | 0 | 3 | 0 | 4 | 3.3 | <0.001 |
| *Meullerius capilaris* | 0 | 0 | 0 | 0 | 1 | 0 | 0 | 1 | 0.8 | 0.432 |
| *Moniezia benedeni* | 0 | 0 | 1 | 0 | 3 | 1 | 0 | 5 | 4.0 | <0.001 |
| *Moniezia expansa* | 0 | 0 | 0 | 0 | 1 | 0 | 0 | 1 | 0.8 | 0.453 |
| *Nanophyetus* sp. | 0 | 0 | 0 | 0 | 0 | 0 | 1 | 1 | 0.8 | <0.001 |
| *Nematodirus* sp. | 0 | 0 | 0 | 0 | 2 | 1 | 0 | 3 | 2.4 | <0.001 |
| *Cryptosporidium* sp. | 0 | 0 | 0 | 0 | 0 | 41 | 1 | 15 | 12.1 | <0.001 |
| *Emeria* sp. | 0 | 0 | 1 | 0 | 0 | 4 | 0 | 5 | 4.0 | <0.001 |
| *Paragonimus kellicot* | 0 | 0 | 0 | 0 | 0 | 2 | 0 | 2 | 1.0 | 0.078 |
| *Strongyloides* sp. | 0 | 0 | 0 | 0 | 0 | 13 | 0 | 13 | 10.6 | <0.001 |
| *Toxocara canis* | 0 | 4 | 0 | 0 | 0 | 0 | 0 | 4 | 4.0 | <0.001 |
| *Toxocara cati* | 1 | 5 | 0 | 0 | 0 | 1 | 0 | 7 | 5.7 | <0.001 |
| *Toxocaris leonina* | 1 | 7 | 0 | 0 | 0 | 0 | 0 | 8 | 6.5 | <0.001 |
| *Trichuris vulpis* | 1 | 0 | 0 | 0 | 1 | 2 | 0 | 4 | 3.3 | <0.001 |
| *Platynosomum fastosus* | 0 | 0 | 1 | 0 | 0 | 0 | 0 | 1 | 0.8 | <0.001 |
| *Isospora* sp. | 0 | 0 | 0 | 1 | 1 | 4 | 0 | 6 | 4.9 | <0.001 |
| **TOTAL** |  |  |  |  |  |  |  | **123** | **100.0** | **<0.001** |

### Prevalence of Parasites among Participants in the Study Sites

**Table 5** shows a parasite prevalence of 67.69% among the study participants: 72.70% in females, 93.80% under ≤ 5 years old, and 25.0% ages≥ 26 years, with at least one parasite in each community, except in Siiroo.

**Table 5:** Prevalence of Parasites among Participants in the Study Sites.

| Variable | Total samples | Positivity Rate | Percentage (%) | X<sup>2</sup> | P-value |
| --- | --- | --- | --- | --- | --- |
| **TOTAL** | **65** | **44** | **67.69%** | **14.892** | **0.0001** |
| **Sex** |  |  |  |  |  |
| Male | 32 | 20 | 62.5 | 0.379 | 0.537 |
| Female | 33 | 24 | 72.7 |  |  |
| **Age** |  |  |  |  |  |
| 0-5 | 32 | 30 | 93.8 | 22.013 | 0.0002 |
| 6-10. | 15 | 6 | 40.0 |  |  |
| 11-15 | 10 | 4 | 40.0 |  |  |
| 16-25 | 4 | 2 | 50.0 |  |  |
| 26 and above | 4 | 1 | 25.0 |  |  |
| **Community** |  |  |  |  |  |
| Berinyasi | 7 | 5 | 71.4 | 4.599 | 0.5961 |
| Ponyenitanga | 3 | 3 | 100 |  |  |
| Siiroo | 1 | 0 | 0.00 |  |  |
| Tanvaare | 12 | 8 | 66.7 |  |  |
| Varinperi | 10 | 6 | 60.0 |  |  |
| Vieri | 28 | 20 | 71.4 |  |  |
| Yeleyiri | 4 | 2 | 50.0 |  |  |

### Parasites recovered from Humans and Domesticated Animals, and the Risk of Zoonosis

**Table 6** shows that parasites such as *Ancylostoma caninum*, host-specific to dogs, were equally recorded in other animals such as cats, ducks, pigs, rabbits, guinea fowls, and humans. Cross-species transmission is attributed to the close intimacy between humans and animals. Also, host-specific parasites to humans, such as *Ascaris* sp., were equally recorded in most of the animals.

**Table 6:** Host-specific parasites shared between humans and domesticated animals.

| Parasite Name | Host(s) | Number of Infections |  | Total Infections | Percentages % |  |
| --- | --- | --- | --- | --- | --- | --- |
|  |  | Human | Animal |  | Human | Animal |
| *Ancylostoma caninum* | Cats, dogs, ducks, guinea fowl, pigs, rabbits, humans. | 1 | 35 | 36 | 2.78 | 97.22 |
| *Ascaris* sp. | Dogs, pigs, humans. | 1 | 6 | 7 | 14.29 | 85.71 |
| *Bunostomum* sp. | Dogs, pigs, humans | 5 | 49 | 54 | 9.26 | 90.74 |
| *Cryptosporidium* sp. | Poultry, rabbits, humans | 5 | 13 | 18 | 27.78 | 72.22 |
| *Eimeria* sp. | Ducks, poultry, rabbits, and humans. | 5 | 78 | 83 | 6.02 | 93.98 |
| *Fasciola* sp. | Cats, humans | 1 | 15 | 16 | 6.25 | 93.75 |
| *Heterakis gallae* | Guinea fowl, poultry, humans. | 1 | 4 | 5 | 20 | 80.00 |
| *Paragonimus kellicotti* | Poultry, humans | 1 | 5 | 6 | 16.67 | 83.33 |
| *Platynosomum fastosus* | Ducks, humans | 3 | 7 | 10 | 30.00 | 70.00 |
| *Strongyloides* sp. | Poultry, humans | 3 | 62 | 65 | 4.62 | 95.38 |
| *Trichuris vulpis* | pigs, poultry, humans | 3 | 13 | 16 | 18.75 | 81.25 |
| *Toxocara canis* | Dogs, goats, humans | 1 | 5 | 6 | 16.67 | 83.33 |
| *Toxocara cati* | Cats, dogs, poultry, humans | 5 | 7 | 12 | 41.67 | 58.33 |
| *Toxocaris leonine* | Cats, dogs, humans | 1 | 8 | 9 | 11.11 | 88.89 |

(Source: Field Data)

## 4. Discussions

This study revealed that the majority (67.0%) of households kept multiple animals, mainly poultry and goats, while over 50.0% had sheep, dogs, and pigs, with a few owning cats and cattle. Most participants in this study reared domestic animals mainly for commercial and livelihood [29]. This practice could be linked to the flagship program of the Government of Ghana on “Rearing for Food and Jobs” [30]. Rearing and owning large animals at home indicates wealth and manhood in this study area [31]. These animals reared at home supplement the family income and serve as a source of protein for the meals at home.

This study demonstrated that backyard farms animals were raised mainly under an extensive system, consistent with previous studies that reported similar rates among humans and domestic animals in Jirel, Nepal [32], and in Ethiopia [33]. This practice is the cheapest for raising domestic animals. However, the practice increases human health risks due to the close interaction between humans and domestic animals, [34], and wild animals as they roam in search of food and water [35]. The animals-human interaction threatens the transmission of infectious agents in the environment from their faecal droppings [18]. In this study, more than 50.0% of the respondents never routinely inspect and monitor the health status of their animals, contrary to over 91.0% reported in Nigeria in which farmers routinely inspected their animals’ wellbeing [36]. This suggests that infected and diseased animals could be reservoirs for potential pathogens and the risks for transmission to humans.

This study reported more than 60.0% parasitic prevalence among seven non-ruminant domesticated animals. This observation was inconsistent with 12.0% and 87.0% previously reported in domestic animals in India [37] and Brazil [38], respectively. This current study also recovered 21 species of gastrointestinal parasites among the non-ruminants, the most prevalent of which were *Ancylostoma caninum*, *Cryptosporidium* sp., and *Strongyloides* sp. This value was much higher than other studies, which reported 11 parasite species in cattle and small ruminants [39] and 13 in dogs [40]. The variations in the parasitic prevalence reported in these studies may be linked to differences in environmental conditions, cultural practices, and animal husbandry systems among households in backyard farms. Dogs, cats, and pigs were most infected in this study. This is consistent with[41] in Brazil and contradicts a previous study in Ghana [42].

In this study, over 67.0% of the human subjects were infected with diverse parasites, consistent with over 60.0% reported among school children with gastrointestinal helminths in Central Mozambique [43] and higher than 15.5% and 20.8% reported in Colombia [44] and Nigeria [45], respectively. These studies reported varied rates associated with the levels of hygienic practices, environmental contamination with animal faeces and socio-cultural practices in the different settings. Most females were infected with parasites, and this observation was in agreement with an earlier report in Yemen et [46]This may be due to limited access to healthcare due to poverty, poor sanitation, and occupational hazards such as caring for children and farming activities.

In this study, most (90%) children under five years were heavily infected. This rate was, however, much higher than 32.81% in Ethiopia [47] and 60.0% in Colombia [44]. The high prevalence in children in this study could be linked to their compromised immune systems, poor personal hygiene, eating food without hand washing, playing on soil contaminated with animal faeces, etc. [48]. Most of the children in this study were infected, which could be attributable to the number of animals in the backyard farms, the kinds of animals reared at home, husbandry and veterinary practices, and the level of animal confinement and human-animal interactions. This study reported large numbers of animals, an extensive system of animal rearing, little or no veterinary care, and high human-animal interactions.

This study recorded 14 parasites in humans out of the 21 species recovered from the non-ruminants in backyard farms, inconsistent with a report in Jirel Nipal [32]. The parasite, *Ancylostoma caninum*, which is host-specific in dogs, was also recovered in cats, ducks, pigs, guinea fowls, and humans. This suggests that cross-species transmission could exist due to close intimacy among the different animal species and humans in the same environment. This is made possible by the different non-ruminants living and sharing the same environment with humans. This cross-species transmission between domestic animals and humans could be attributed to the extensive animal farming system, limited veterinary care, poor sanitation and environmental practices, poor housing [49] and inadequate knowledge of how zoonoses are transmitted [9]. Hence, there is an urgent need to implement the One-Health Concept to avert possible zoonosis outbreaks. No significant difference was recorded between the prevalence of parasitic zoonosis in humans and domestic animals, consistent with an earlier study in Egypt [50].

This study also noted that parasites such as *Ancylostoma caninum, Toxocara canis, Toxocaris leonina, and Trichuris vulpis*, whose specific hosts are dogs, were recovered in other animals and humans. *Bunostomum* sp., *Fasciola* sp*., Haemonchus contortus, Eimeria* sp*., Schistosoma bovis,* and *Strongylus* sp., whose specific hosts are ruminants, were isolated in other non-ruminants including humans. *Cryptosporidium s*p*., Ascaris* sp*., Paragonimus kellicotti,* and *Strongyloides* sp., whose specific hosts are humans recovered in non-ruminant domestic animals [32], and this supports an earlier report in Nigeria [45]. *Toxocara cati* and *Platynosomum fastosum*, whose hosts are cats, were recovered in humans and other animals in this study area. These findings further highlight the human-animal-animal-specific species cross-transmission of the various parasites, supporting the One-Health Concept. This poses dire public health consequences and implications for human and animal health [51]. This study is a novelty linking animal-to-animal host-specific species transmission and human-to-animal host-specific parasite species cross-transmission. The findings in this study call for an urgent need for stakeholders to dialogue and roll out the implementation of the One-Health Concept for the welfare of humans and animals alike and to forestall the rampant outbreaks and epidemics of emerging and reemerging diseases linked to domestic and wife life animals.

It has been noted that the cross-species transmission of zoonotic parasites, which are of public health importance, further demonstrated the interconnectedness of human, animal, and environmental health [52]. Therefore, it is prudent to foster collaborations between different sectors to maintain the well-being of humans, animals, and the environment by employing the principles of the One-Health Concept to mitigate the risk of zoonosis transmission [53].

### Study Limitations

Several setbacks were encountered during this study. Firstly, some participants within the study communities were unwilling to participate due to superstitious beliefs, accounting for the relatively small stool sample size for humans. However, absolute confidentiality was assured to them. Also, most of the participants were illiterates, and extra time was needed to interpret the questionnaire items in their local dialect. Isolation and identification of the different parasites were challenged, and molecular techniques could make this more accurate and specific.

## 5. Conclusion

The study reveals a high prevalence of parasites among non-ruminants and humans of zoonotic origin in the Wa West District. This study recovered 21 parasites of zoonotic importance among non-ruminants, of which 14 were recorded in both humans and domestic animals. *Ancylostoma caninum*, *Cryptosporidium* sp., and *Strongyloides* sp. were the most prevalent parasites recovered from non-ruminants and humans. The study also noted that host-specific parasites such as *Ancylostoma caninum, Toxocara canis, Toxocaris leonina, and Trichuris vulpis* in dogs were recovered in other animals and humans due to the close relationships within a limited space in each household. This has dire implications for animal and human health and calls for implementing the One-Health Concept. It is recommended that an effective collaboration between the Ghana Health Service, Animal Health Service, and the Environmental Health Service foster and combat the possible outbreak of zoonosis within the study.

## Data Availability

All data used for this manuscript are available upon a reasonable request.

## Acknowledgements

This study was approved by the Committee on Human Research, Publications, and Ethics at the Kwame Nkrumah University of Science and Technology (REF: CHRPE/AP/851/23) and the Ghana Health Service at the Wa West District Assembly. It was conducted in partial fulfillment of the requirements for the Master of Philosophy degree in Biology at AAMUSTED.

We sincerely appreciate Mr. Shaibu Waale and Mr. Sixtus Saa-Ere for their invaluable laboratory contributions. We also extend our gratitude to the District Chief Executive (DCE) and the staff of the Wa West District Assembly for their support and permission to conduct this study within their jurisdiction.

## Availability of supporting data

All data used for this manuscript are available upon a reasonable request.

## Authors Contributions

JKZ was responsible for data collection, data analysis, and manuscript drafting. DDY played a pivotal role in conceptualizing and developing the manuscript, provided critical edits, contributed to data analysis and interpretation, and ensured the manuscript’s intellectual rigor and clarity. NYAB reviewed the manuscript and approved it for publication. EA managed the data, reviewed the manuscript, approved it for publication and prepared the manuscript for submission. All authors have thoroughly read and approved the final manuscript for publication.

## Consent for publication

We have all read the final manuscript and consented to it for publication.

## References

1. Rahman, M.T., et al., Zoonotic diseases: etiology, impact, and control. 2020. 8(9): p. 1405.

2. Abebe, E., G. Gugsa, and M.J.J.o.t.m. Ahmed, Review on major food-borne zoonotic bacterial pathogens. 2020. 2020(1): p. 4674235.

3. Dharmarajan, G., et al., The animal origin of major human infectious diseases: what can past epidemics teach us about preventing the next pandemic? 2022. 2(1).

4. Sohail, M., et al., The Threat of Transboundary Zoonosis. 2023. 4: p. 701–715.

5. Tabish, S.A. and S.J.H. Nabil, An age of emerging and reemerging pandemic threats. 2022. 14(10): p. 1021–1037.

6. Thoradeniya, T., S.J.G. Jayasinghe, and health, COVID-19 and future pandemics: a global systems approach and relevance to SDGs. 2021. 17(1): p. 59.

7. Edward, M., et al., Climate change and contagion: the emerging threat of zoonotic diseases in Africa. 2025. 15(1): p. 2441534.

8. Dixon, J., et al., Farming systems and food security in Africa. 2020.

9. Esposito, M.M., et al., The impact of human activities on zoonotic infection transmissions. 2023. 13(10): p. 1646.

10. Caminade, C., K.M. McIntyre, and A.E.J.A.o.t.N.Y.A.o.S. Jones, Impact of recent and future climate change on vector-borne diseases. 2019. 1436(1): p. 157–173.

11. Asante, J., A. Noreddin, and M.E.J.P. El Zowalaty, Systematic review of important bacterial zoonoses in Africa in the last decade in light of the ‘One Health’concept. 2019. 8(2): p. 50.

12. Ellwanger, J.H., J.A.B.J.G. Chies, and M. Biology, Zoonotic spillover: Understanding basic aspects for better prevention. 2021. 44: p. e20200355.

13. Efua, A.J.J.o.A.H., Role of Wildlife in The Transmission of Zoonotic Diseases in Ghana. 2023. 3(1): p. 45–56.

14. Harl, J., et al., Novel phylogenetic clade of avian Haemoproteus parasites (Haemosporida, Haemoproteidae) from Accipitridae raptors, with description of a new Haemoproteus species. 2024. 31.

15. Amoako, Y., et al., Rabies is still a fatal but neglected disease: a case report. 2021. 15: p. 1–6.

16. Saaka, M. and J.J.S. Akuamoah-Boateng, Prevalence and determinants of rural-urban utilization of skilled delivery services in Northern Ghana. 2020. 2020.

17. Edao, B.M., et al., Brucellosis in ruminants and pastoralists in Borena, Southern Ethiopia. 2020. 14(7): p. e0008461.

18. Marrana, M., Epidemiology of disease through the interactions between humans, domestic animals, and wildlife, in One Health. 2022, Elsevier. p. 73–111.

19. Issah, B., T. Ansah, and H.J.U.I.J.o.D. Alagma, Awareness of zoonotic diseases among pet owners in wa municipality of ghana. 2020. 7(2): p. 387–397.

20. Ansah-Owusu, J., et al., Tick-borne pathogens of zoonotic and veterinary importance in cattle ticks in Ghana. 2024. 123(1): p. 44.

21. Amoah, L., Potential Transmission Of Zoonoses Between The Human-Domestic-Wildlife Interface And Its Implication For Sustainable Health. 2021, University Of Ghana.

22. GhanaWeb, Wa West is a ‘zoo’ of diseases – District Health Director. 2018.

23. Gwenzi, W., et al., Grappling with (re)-emerging infectious zoonoses: Risk assessment, mitigation framework, and future directions. 2022. 82: p. 103350.

24. Amoah, J.O., J.J.W. Mensah, Space, and Society, Gender and public works intervention in rural Ghana: An empowerment framework perspective. 2023. 5: p. 100176.

25. Abdulai, I.A., A. Dongzagla, and A.J.U.G. Ahmed, Urban livestock rearing and the paradox of sustainable cities and urban governance in West Africa: Empirical evidence from Wa, Ghana. 2023. 3(4): p. 304–314.

26. Sulieman, Y., M.A. Zakaria, and T.J.A.o.p. Pengsakul, Prevalence of intestinal helminth parasites of stray dogs in Shendi area, Sudan. 2020. 66(1).

27. Silva, V., et al., Epidemiological survey on intestinal helminths of stray dogs in Guimarães, Portugal. 2020. 44(4): p. 869–876.

28. Mulu, W., et al., Campylobacter occurrence and antimicrobial resistance profile in under five-year-old diarrheal children, backyard farm animals, and companion pets. 2024. 18(6): p. e0012241.

29. Mukhtar, A., B.Z. Abdulkarim, and T.A.J.I.J.H.S.S. Ladan, Socio-Economic Benefits of Livestock Rearing In Maiduguri Metropolis, Borno State, Nigeria. 2021. 26(1): p. 16–23.

30. Asante, F.A. and S. Bawakyillenuo, Farm-level effects of the 2019 Ghana planting for food and jobs program: An analysis of household survey data. Vol. 57. 2021: Intl Food Policy Res Inst.

31. Craighead, L., et al., “Everything in this world has been given to us from cows”, a qualitative study on farmers’ perceptions of keeping dairy cattle in Senegal and implications for disease control and healthcare delivery. 2021. 16(2): p. e0247644.

32. Dhakal, P., et al., Prevalence of intestinal parasites in humans and domestic animals in Jirel community, Dolakha, Nepal. 2024. 13(8): p. 3408–3414.

33. Ayelign, A. and T.J.H. Zerfu, Household, dietary and healthcare factors predicting childhood stunting in Ethiopia. 2021. 7(4).

34. Silva, S.R., et al., Extensive sheep and goat production: The role of novel technologies towards sustainability and animal welfare. 2022. 12(7): p. 885.

35. Abdulai, I.A.J.H., Rearing livestock on the edge of secondary cities: examining small ruminant production on the fringes of Wa, Ghana. 2022. 8(4).

36. Eniola, O., et al., Perception and safety practices to zoonotic diseases transmission among small ruminant farmers in ona-ara local government area of oyo state. 2020. 12(2).

37. Malathi, S., U. Shameem, and M.J.J.o.P.D. Komali, Prevalence of gastrointestinal helminth parasites in domestic ruminants from Srikakulam district, Andhra Pradesh, India. 2021. 45(3): p. 823–830.

38. Zanetti, A.S., et al., Diversity and prevalence of intestinal parasites of zoonotic potential in animal hosts from different biomes in the central region of Brazil. 2021. 67(1).

39. Squire, S., et al., Prevalence and risk factors associated with gastrointestinal parasites in ruminant livestock in the Coastal Savannah zone of Ghana. 2019. 199: p. 105126.

40. Amissah-Reynolds, P.K., et al., Prevalence of helminths in dogs and owners’ awareness of zoonotic diseases in Mampong, Ashanti, Ghana. 2016. 2016.

41. Souza, J.B.B., et al., Prevalence of intestinal parasites, risk factors and zoonotic aspects in dog and cat populations from Goiás, Brazil. 2023. 10(8): p. 492.

42. Amissah-Reynolds, P.K.J.I.J.P.R., Zoonotic risks from domestic animals in Ghana. 2020. 4: p. 17–31.

43. Muadica, A., et al., Risk associations for intestinal parasites in symptomatic and asymptomatic schoolchildren in central Mozambique. 2021. 27(4): p. 624–629.

44. Peña-Quistial, M.G., et al., Prevalence and associated risk factors of Intestinal parasites in rural high-mountain communities of the Valle del Cauca—Colombia. 2020. 14(10): p. e0008734.

45. Obebe, O.O., Intestinal parasitic infections in HIV-infected patients and domestic animals in Ekiti State, Nigeria. 2024.

46. Edrees, W.H., M.A. Alshahethi, and M.S.J.A.-R.U.J.f.M.S. Al-Awar, Risk Factors Associated with Prevalence of Intestinal Parasitic Infections among Schoolchildren in Amran City, Yemen. 2022. 6(2).

47. Amare, Y., T. Yohannes, and S.J.J.o.P.D. Tesfaye, A cross-sectional study on the prevalence and associated risk factors for intestinal parasitic infections among under-five aged children in Dabat primary hospital, north Gondar, Ethiopia. 2024: p. 1–13.

48. Domenico, B., et al., The impact of environmental alterations on human microbiota and infectious diseases, in Environmental Alteration Leads to Human Disease: A Planetary Health Approach. 2022, Springer. p. 209–227.

49. Schatz, A.M. and A.W.J.P.o.t.R.S.B. Park, Host and parasite traits predict cross-species parasite acquisition by introduced mammals. 2021. 288(1950): p. 20210341.

50. Shehab, A.Y., et al., Intestinal parasites among humans and their livestock animals in a rural community in Gharbia governorate, Egypt. 2021. 45: p. 96–100.

51. Achi, C.R., et al., Operationalising One Health in Nigeria: reflections from a high-level expert panel discussion commemorating the 2020 World Antibiotics Awareness Week. 2021. 9: p. 673504.

52. Erkyihun, G.A. and M.B.J.Z. Alemayehu, One Health approach for the control of zoonotic diseases. 2022. 2(1): p. 963.

53. Ranganatha, S., et al., “One health” concept to mitigate zoonoses: Indian perspective. 2023.

